# SARS-CoV-2 genome surveillance in Mainz, Germany, reveals convergent origin of the N501Y spike mutation in a hospital setting

**DOI:** 10.1101/2021.02.11.21251324

**Authors:** NA Lemmermann, B Lieb, T Laufs, A Renzaho, S Runkel, W Kohnen, M Linke, S Gerber, S Schweiger, A Michel, S-E Bikar, B Plachter, T Hankeln

**Author notes:** correspondence to NL, Niels A. Lemmermann, University Medical Center Mainz, Institute for Virology, Obere Zahlbacher Str. 67, 55131 Mainz, Germany. joint senior authors (S-E Bikar, B Plachter, T Hankeln). equal contribution (NA Lemmermann, B Lieb).

## Abstract

While establishing a regional SARS-Cov-2 variant surveillance by genome sequencing, we have identified three infected individuals in a clinical setting (two long-term hospitalized patients and a nurse) that shared the spike N501Y mutation within a genotype background distinct from the current viral variants of concern. We suggest that the adaptive N501Y mutation, known to increase SARS-CoV-2 transmissibility, arose by convergent evolution around December in Mainz, Germany. Hospitalized patients with a compromised immune system may be a potential source of novel viral variants, which calls for monitoring viral evolution by genome sequencing in clinical settings.

## Introduction

Coronaviruses are (+)ssRNA viruses with a moderate to high mutation rate compared to other ssRNA viruses and a calculated substitution rate of about 10^−3^ to 10^−4^ per nucleotide and year (Su et al. 2016; Zhao et al. 2004). Severe acute respiratory syndrome coronavirus 2 (SARS-CoV-2), the beta-coronavirus that causes the ongoing pandemic with substantial morbidity and mortality worldwide, displays a mutation rate in the same order of magnitude (Coronavirus Study Group, 2020; Wu et al. 2020, Zhou et al. 2020). However, probably due to the very large population size of this virus with phases of exponential growth (Moya et al. 2004; Vasilarou et al. 2020), thousands of changes in the genome have already been described. While most of them will not affect viral properties, natural selection is highly efficient in such large populations, and adaptive mutations that increase viral fitness are likely to occur (Moya et al. 2004). In fact, several SARS-CoV-2 variants have already emerged during the pandemic, showing enhanced host adaptation associated with increased infectivity and transmission. All of these variants show specific mutations in the S gene which encodes the spike (S) protein. The S protein mediates binding of the virus to the cellular angiotensin-converting enzyme 2 (ACE2) receptor and subsequent fusion with the host cell. Therefore, the S protein is the target for vaccines as well as therapeutic antibody development. The first evolutionarily successful mutation of the S protein, D614G, was initially observed in China in March 2020 and rapidly spread worldwide until the end of 2020 (Korber et al. 2020; Yurkovetskiy et al. 2020). In September and December 2020, additional virus variants with conspicuous S mutations have been reported (the latter three have also been designated as *variants of concern*, VOC): The “cluster 5/mink” variant isolated in Denmark is marked by the S gene ΔH69/ΔV70 deletion in association with the receptor binding domain (RBD) mutation Y453F (ECDC 2020). The B1.1.7 VOC (*syn*. 20I/501Y.V1) possibly emerged in the UK and is now rapidly spreading across the country and into continental Europe (Rambaut et al. 2020). It carries multiple changes in addition to ΔH69/ΔV70, among which the RBD mutations N501Y and P681H are the most notable ones. In the VOC B.1.351 (*syn*. 20H/501Y.V2), spreading recently in South Africa, the N501Y mutation is combined with the replacements K417N and E484K, all three affecting the RBD (Tegally et al. 2020). Another rapidly spreading mutant virus lineage possibly originating in Brazil, B.1.1.28 (P1; *syn*. 20J/501Y.V3), also carries these three changes, but with the variation K417T (Vasques Nonaka et al. 2020).

Structural analyses of the RBD provided evidence for the importance of N501 and K417 in the interaction of the S protein with ACE2 (Lan et al. 2020, Shang et al. 2020). N501 is part of the receptor binding motif (RBM), which includes most of the direct interacting residues, and forms a hydrogen bond to the Y41 of ACE2 that is further stabilized by the E484 residue. K417 is a unique ACE2-interacting residue outside of the RBM. Amino acid changes N501Y and E484K enhance the binding affinity of the S protein to ACE2 which might be correlated to the epidemiologically observed higher infectivity and transmission (Zahradnik et al. 2021). Furthermore, both exchanges are discussed as a potential risk for immune escape (Nelson et al. 2021, Wibmer et al. 2021).

Monitoring the mutational spectrum that emerges within the S gene of SARS-CoV-2 is thus most critical for the implementation of interventional methods and combatting the pandemic. The origin and evolution of specific mutations also deserve attention. While variant D614G clearly originated once and then spread worldwide, the divergent geographic distribution of the three new virus lineages which are jointly characterized by the N501Y mutation suggests an independent origin, implying convergent evolution. Will N501Y mutations thus independently arise more often in the future, and under which circumstances? The mutation harbors the potential for virus escape, especially in immunocompromised patients presenting with prolonged viral replication. Also, therapeutic intervention with neutralizing antibodies could be a scenario that enhances S protein-mediated evasion and the outgrowth of escape variants (Williams & Burgers, 2021).

Initiating coordinated efforts for regional SARS-CoV-2 variant surveillance by genome sequencing, we detected a micro-cluster of three individuals in a hospital setting, where N501Y was reproducibly found, but without any other typical mutations of the known variants of concern. The scenario strongly suggests the possibility of a convergently recurring emergence of critical SARS-CoV-2 spike mutations.

## Materials and Methods

### Sample acquisition, RNA isolation and PCR testing

Nasopharyngal swabs were taken routinely for SARS-CoV-2 diagnostics by health care professionals, eluted in Virocult transport medium (MWE, Corsham, GB) and analyzed by RT-qPCR for SARS-CoV-2 RNA within 18h. RNA isolation and RT-qPCR were performed for the N- and Nsp-2 gene on a NeuMoDx 288 Molecular System (Molecular Systems, Ann Arbor, USA). A full list of the studied samples including sample type and origin, PCR Ct values and genome sequencing parameters is provided in Tab. 1.

### PCR validation for N501Y

The presence of the N501Y mutation was identified in the isolates by melting curve analysis, using the RTVirSNiP SARS-CoV-2 Spike N501Y Kit (TibMolbiol, No. 53-0780-96, Berlin, Germany) according to the supplier’s instructions. Briefly, viral RNA was isolated from stored nasopharyngeal swabs using the EMAG system (Biomerieux, Nürtingen, Germany). Purified viral RNAs were subsequently deployed to identify the nucleotide substitution A23063T (corresponding to N501Y) in the S gene. To this end, a 130 Bp fragment was amplified and analyzed by melting curve assay on a 7500 Real-Time PCR System (Applied Biosystems, Darmstadt, Germany).

### Whole-genome sequencing and bioinformatic processing

Sars-CoV-2 genome sequences were inferred using a targeted amplicon-based strategy. First, purified viral RNA was converted to cDNA by random hexamer priming using the Lunascript RT kit (New England Biolabs, Frankfurt, Germany). Typically, 5-10 ul of the purified RNA extract were used for reverse-transcription. Additionally, a negative control reaction was set up, containing water instead of RNA extract. We then applied the EasySeq RC-PCR SARS CoV-2 Whole Genome Sequencing Kit (NimaGen, Nijmegen, NL) for amplifying the whole viral genome by tiled PCR primer sets, which are based on the ARTIC standard protocol (https://artic.network/ncov-2019). Libraries were inspected on a QIAxcel capillary electrophoresis (Qiagen, Hilden, Germany), normalized to about 9 pM and run on a MiSeq sequencer (Illumina, Berlin, Germany; 2×250 bp v2 chemistry; 2×150 bp for samples #19, #20, #21, #22, #24, #25). Between 500 k and 1.5 mio reads were received per sample.

For bench-marking purposes, we applied two bioinformatic analysis pipelines in parallel, the virSEAK pipeline (JSI medical systems, Ettenheim, Germany) and the Geneious prime platform (Biomatters, Auckland, NZ). Sequence reads were processed by quality trimming, removal of Illumina adapters, and amplicon-specific PCR primers. Thereafter, amplicon reads were mapped to the SARS-CoV-2 reference sequence Wuhan-Hu-1 (NC_045512), followed by calling of the sequence variants. Read depth typically ranged between 3000 and 6000 per nucleotide position. Both pipelines essentially produced the identical set of single nucleotide variants (SNVs) for the studied genomes. In addition to automatic calling, SNVs were inspected visually by the SeqNext (JSI medical systems) and Geneious prime tools, respectively. Variants were typically called using a coverage threshold of 20 reads and a frequency of at least 90%. Thereafter, variants were integrated into the reference sequence to produce consensus sequence files in FASTA format for each individual genome. FASTA genome consensus sequences were submitted electronically to the GISAID repository (www.gisaid.org) and to the Robert Koch Institute, Germany (www.rki.de).

Initial lineage assignment for all genomes was performed by applying the NextClade v0.12.0 online tool (https://clades.nextstrain.org), virSEAK (JSI medical systems) and Pangolin (https://pangolin.cog-uk.io/). For phylogenetic tree reconstruction, individual FASTA genome files were combined into a single multi-sequence FASTA file and aligned by MAFFT v7.450 (Katoh and Standley 2013, Katoh et al. 2002) using default criteria. Maximum likelihood trees were calculated using RAxML (Stamatakis 2014) under the GTR substitution model and gamma distribution with 1000 bootstraps. Trees were visualized in Geneious prime, showing bootstrap support values at the respective nodes.

Mapping and visualization of individual amino acid replacements in the SARS-CoV-2 spike protein (data not shown) were produced using the CoVsurver online tool (https://www.gisaid.org/epiflu-applications/covsurver-mutations-app/).

## Results

While establishing a regional SARS-CoV-2 surveillance routine for the State of Rhineland-Palatinate, the South-West of Germny and the city of Mainz, we sequenced the genomes of 25 samples from 20 individuals, obtained from clinical settings at the University Medical Center Mainz during December 2020 and early January 2021 (Tab. 1). The samples were tested positive for SARS-CoV-2 RNA by routine RT-PCR for the N and NSP2 genes. PCR-positive samples with Ct values ranging from 10 to 27 yielded complete genome sequences with a mean read depth (coverage) ranging between 2,000 and 13,700. Between 16 and 29 variant positions were identified per genome in comparison to the reference sequence (Fig. 1, Tab. 1). According to the Nextstrain classification (https://nextstrain.org/blog/2021-01-06-updated-SARS-CoV-2-clade-naming), samples could be assigned to different viral clades, i. e. to the globally distributed clades 20B (n=7) and 20A.EU2 (n=2), to clade 20E.EU1 (n=14), probably of European origin, or to clade 20A (n=1) (Tab. 1; Fig. 2). All genomes shared the spike protein mutation D614G, which has spread worldwide since springtime 2020. To infer relationships and possible transmission routes, the Mainz genome sequences were analyzed by phylogenetic tree reconstruction (Fig. 3). Results indicated one major cluster (bootstrap support 100%) containing 14 genomes, which in the clinical setting derived from the same ward. Within this cluster, some sequences were even almost identical, arguing in favour of potential direct transmission events. On a separate branch (bootstrap support 100%), we identified a micro-cluster of replicate samples from two patients who displayed an identical SARS-CoV-2 genomic sequence (samples #4, #23). These two individuals had a negative PCR test result upon hospital admission, were hospitalized for ca. 2 weeks and shared the same room, but resided on a different ward than the other patients in the cohort. Their sequences were characterized by a long branch in the phylogenetic tree, indicative of fast sequence evolution (Fig. 3).

**Fig. 1.**
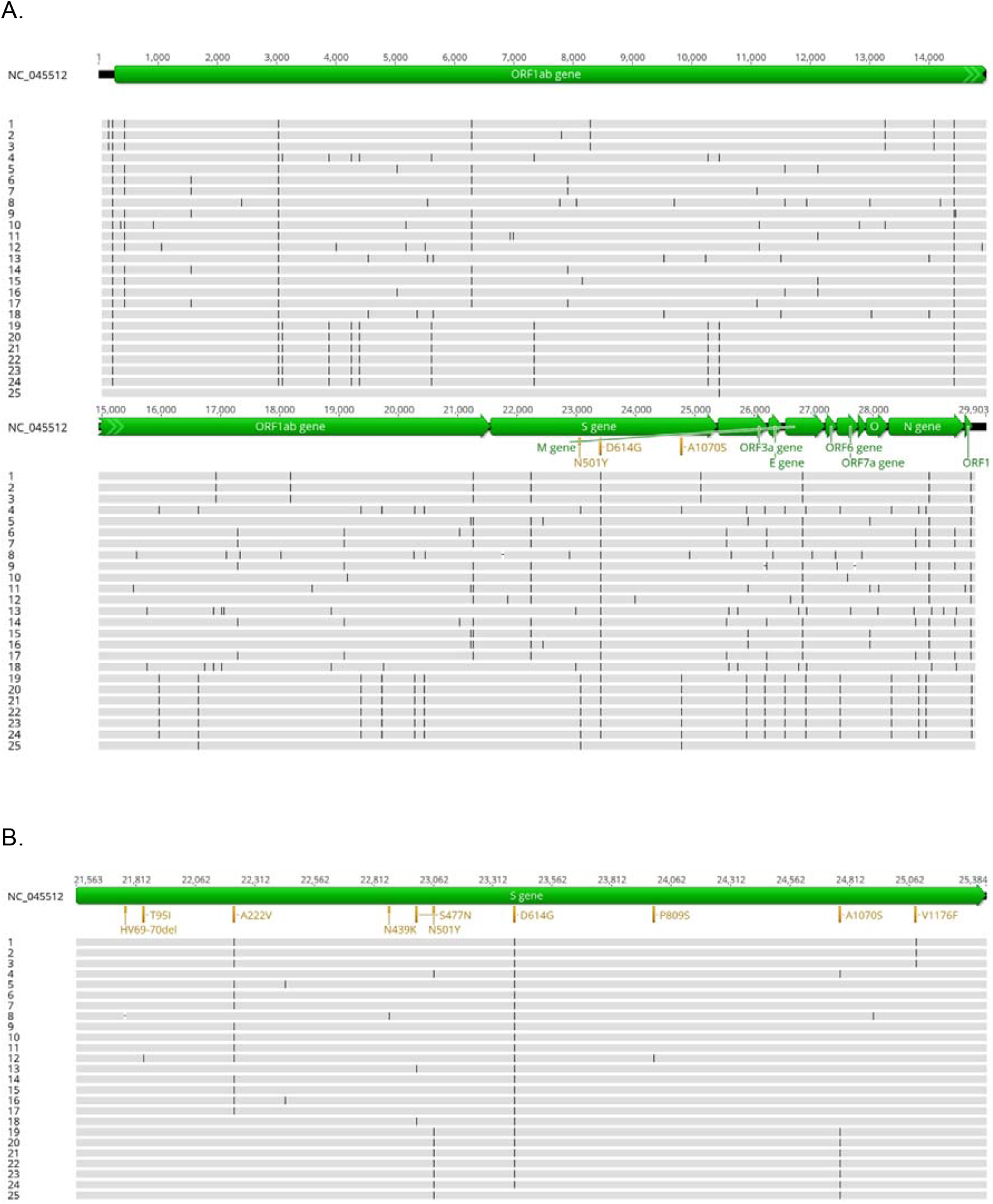
Position of mutations observed in the sequenced SARS-CoV-2 samples, shown for the full genomes (A) and enlarged for the S gene (B). Sample names given at the right side correspond to Tab. 1.

**Fig. 2.**
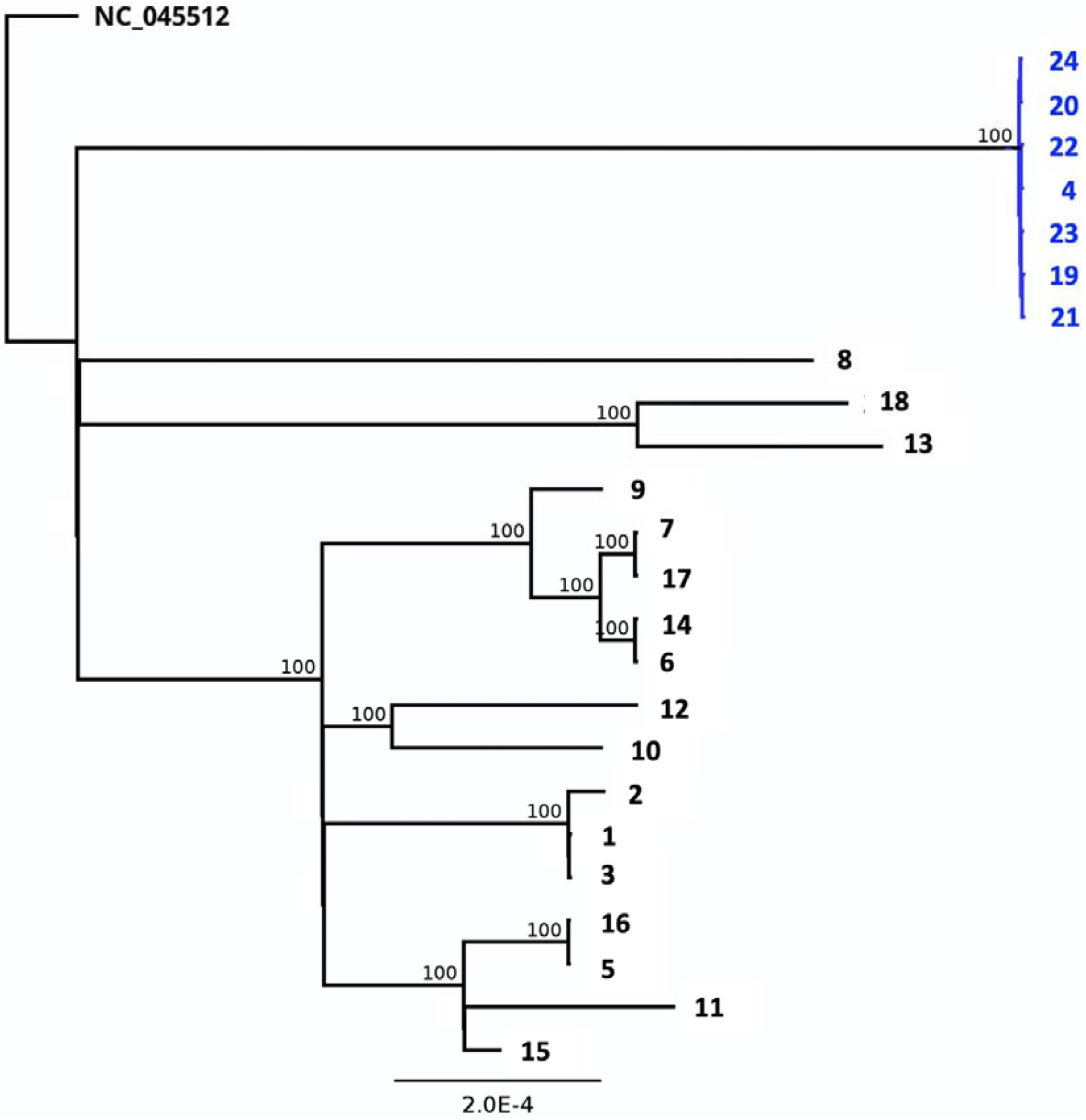
Maximum likelihood phylogenetic tree based on full genome sequences reveals the relationships of the study samples. Bootstrap values are indicated at the respective nodes. The cluster of sequences labeled in blue originates from the samples of the two patients and contains the N501Y spike mutation. Sample names at the branch tips correspond to Tab. 1.

**Fig. 3.**
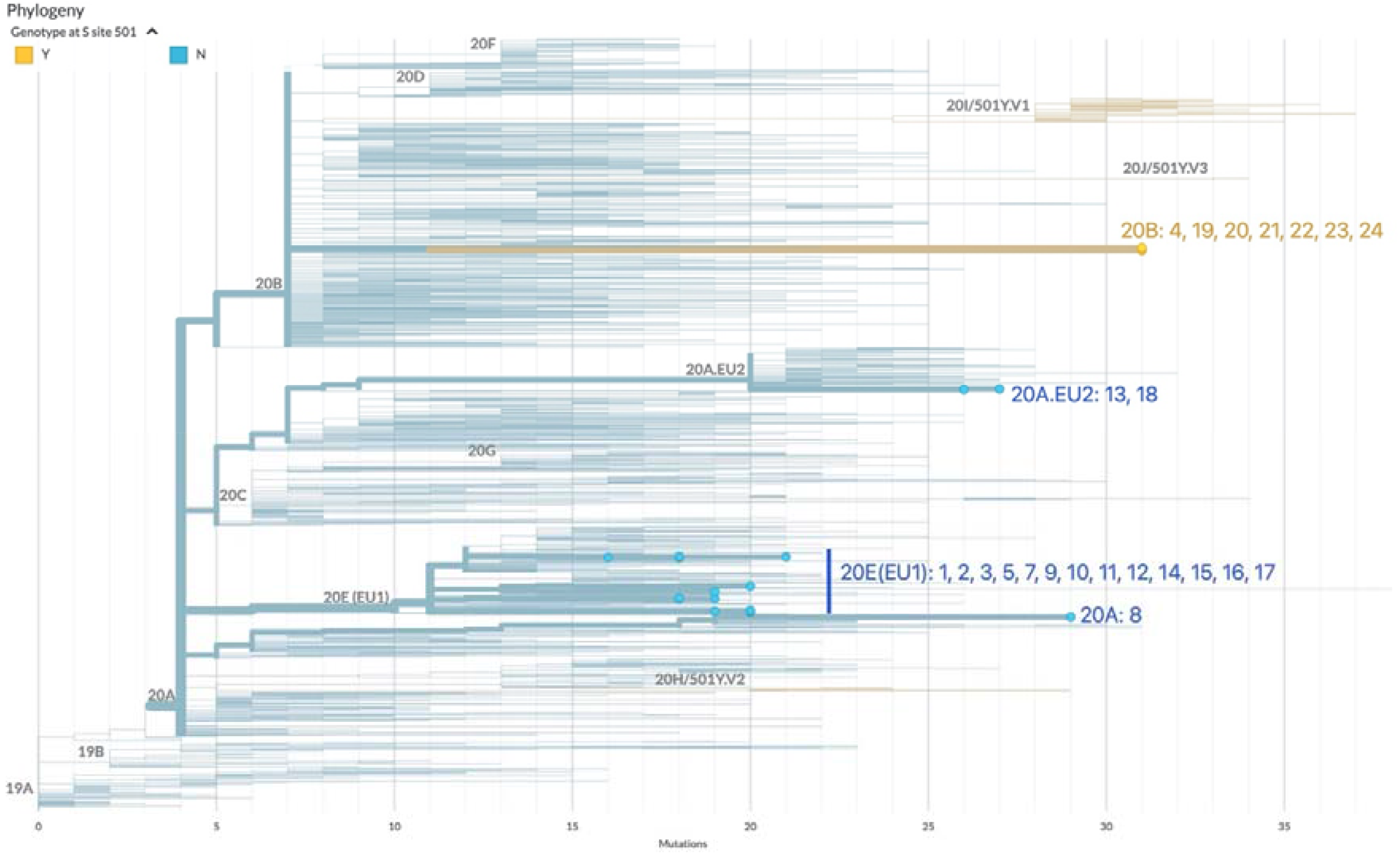
Clade classification of the study sample according to “Nextstrain”. Major clades are indicated along the tree branches. Study samples are depicted with coloured tips (blue, yellow), their designations correspond to Tab. 1. The position of the currently known VOCs is also indicated.

**Tab. 1.**
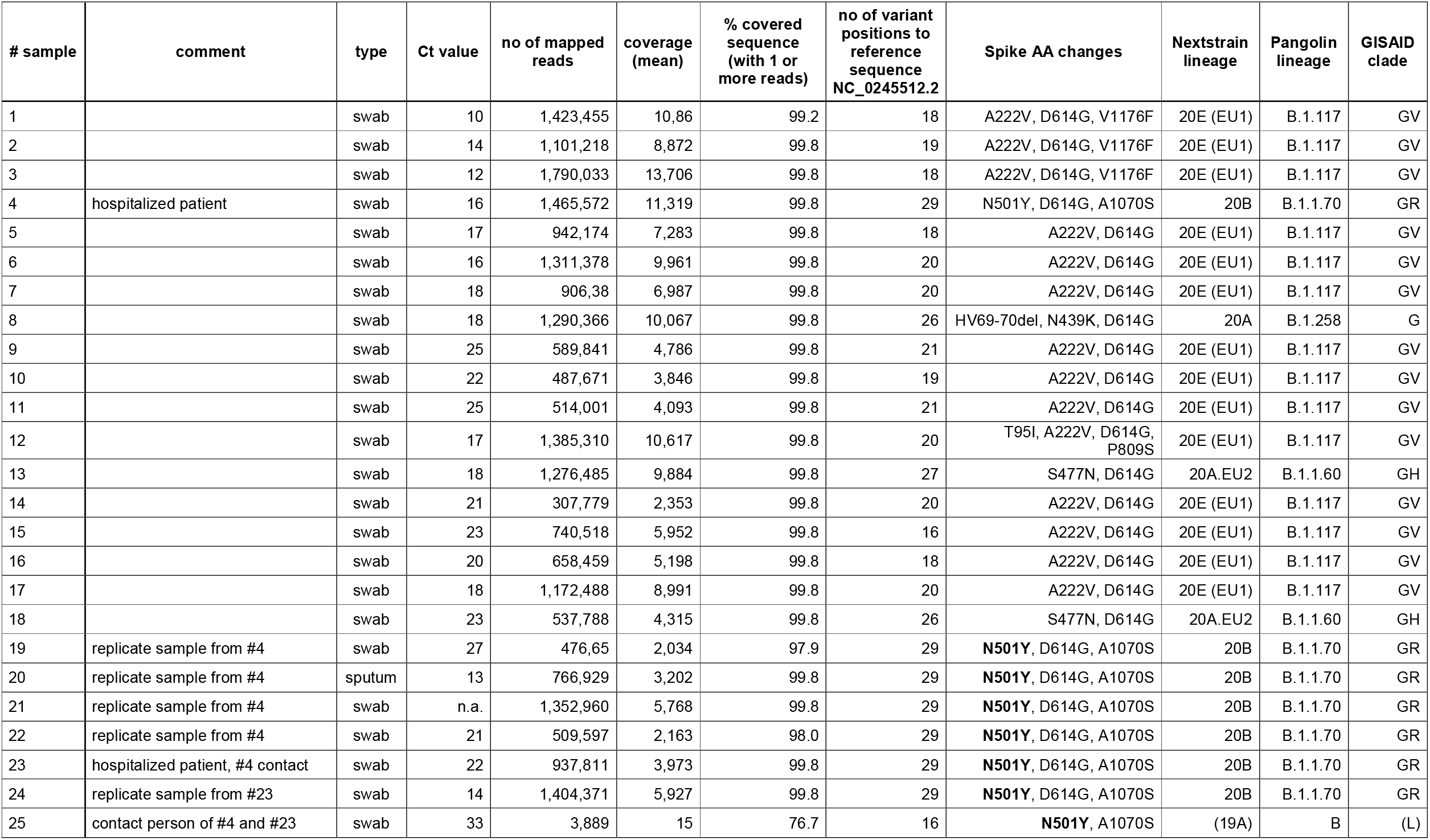
Study sample overview

The subsequent detailed inspection of the identified genomic sequence variants from the full study cohort showed that neither sample belonged to the three virus lineages currently creating the most concern internationally, i. e. VOC B.1.1.7, B.1.351 and B.1.1.248 (P.1). Interestingly, however, in the samples collected from the two room-neighbor patients in December 2020 (#4, #23) we could unambiguously identify the spike receptor binding domain (RBD) mutation N501Y in combination with another change in the S protein, A1070S. The latter is relatively rare with a frequency of 0.034 in the GISAID genome database (www.gisaid.org) and resides in the spike S2 subunit which is involved in viral membrane fusion and cell entry. None of the other spike changes characteristic for VOC lineages B.1.1.7, B.1.351, and B.1.1.248 could be detected. This result strongly suggests that the N501Y change present in samples #4 and #23 has evolved independently by convergence in a clade 20B genotype background in December in Mainz, Germany. Both individuals did not have a travel history.

We confirmed the genotypes of both, #4 and #23, by sequencing additional RNA samples taken from the very same patients at earlier or later dates in December. We did neither observe fixed genetic differences between the samples, nor sequence heterogeneity within samples, possibly indicative of ongoing viral evolution. In addition, the N501Y mutation was confirmed by RT-PCR and subsequent melting curve analysis (Fig. 4). Looking for additional contact individuals of the two patients, material was sequenced from a nurse working on the same ward (#25), who had tested PCR-positive around the same time. Due to a high PCR cycle threshold (Ct of 33) and thus very little RNA material, only a few thousand reads were obtained for this person during sequencing and no full-length genome sequence could be reconstructed (Tab. 1). However, positions N501Y and A1070S could also be confirmed in this individual.

**Fig. 4.**
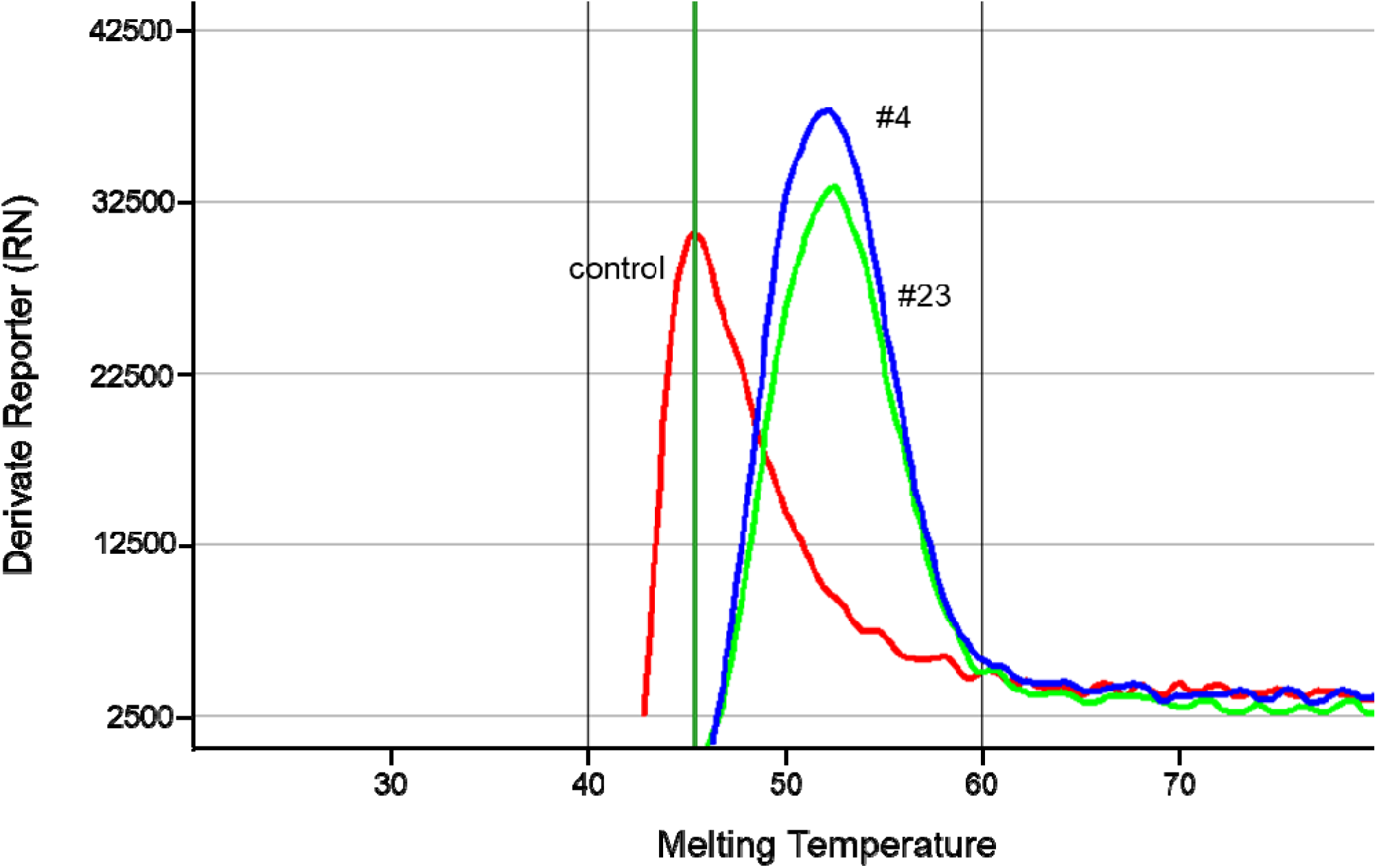
Confirmation of the N501Y mutation in samples #4 and #23 by PCR and subsequent melting curve analysis.

## Discussion

In early 2021, mutant viral lineages designated B.1.1.7, B.1.351 and B.1.1.248 (P.1), each having a unique combination of mutations in their spike (S) gene, are creating much concern due to their presumed higher transmissibility and potential for immune system escape, illustrating the key role of variant surveillance by genome sequencing (Cyranoski 2021, Mahase 2021). A mutation which is common to the S genes of these three virus variants is resulting in the N501Y amino acid change within the RBD of the S protein. N501Y substantially increases the binding affinity of S to its cellular receptor ACE2, thus ameliorating virus entry (Starr et al. 2020; Zahradnik et al. 2021). In the viral VOCs mentioned above, N501Y is accompanied by additional lineage-specific mutations that may render the viral phenotype even more problematic (Zahradnik et al. 2021).

The divergent geographic origin of those VOCs makes it likely that they emerged in separate events, possibly in late 2020, suggesting that the mutual N501Y change evolved in parallel by convergence. Here we report the detection of the same N501Y mutation in a small cluster of individuals, who were tested positive for SARS-CoV-2 in a clinical setting in Mainz, Germany. We found N501Y in a genomic background distinct from the known VOCs, thus occurring out of the context of their additional S mutations. This lends further evidence to the hypothesis of convergent evolution of the N501Y change. In fact, a genome surveillance group based in Ohio, USA, very recently also reported such local emergence of N501Y from a still different viral clade, 20C (Tu et al. 2021). This scenario suggests that the N501Y mutation is an initial key event in driving the evolutionary success of SARS-CoV-2, and its emergence should be monitored world-wide with attention. Preliminary data now indicate that such an initial N501Y mutation can – probably anytime and anywhere -be followed by a second, phenotype-aggravating event: in mid-January 2021, the E484K spike mutation, previously reported to enhance infectivity specifically in the “South-African” B1.351 and the “Brasilian” B.1.1.248 (P1) VOCs (Tegally et al. 2020; Faria et al. 2021), was first-time detected as a novel feature in several samples of the “British” VOC B.1.17 and thus probably also emerged within this lineage by convergence (PHE 2021).

But under which circumstances do these spike gene mutations arise in humans, and which conditions favor their adaptive selection? In our particular hospital micro-cluster with the two bed neighbour patients and one infected nurse, the definite origin of the N501Y spike mutation cannot be inferred unambiguously. Further cases carrying this exact genotype have not been identified so far, and the SARS-CoV-2 cases in the neighboring ward had different roots (comp. Fig. 1). However, we wish to point out that the two Mainz patients both were immunocompromised and hospitalized for ca. 2 weeks. In fact, Kemp et al. (2020) and Chio et al. (2020) recently reported individual cases of immunocompromised patients in whom SARS-CoV-2 variants evolved rapidly during long-term infection, showing elevated nucleotide substitution rates and the emergence of conspicuous S amino acid changes (including N501Y in one case).

In consequence, we suggest to focus some attention during the future SARS-CoV-2 surveillance by genome sequencing on clinical settings which might favor viral immune escape and the selection of mutations of clinical relevance. While PCR testing (e. g. specifically for the N501Y or E484K mutations) can readily be applied for rapid pre-screening, only whole-genome sequencing will uncover the full repertoire of changes that determine the evolution and clinical phenotype of this virus.

## Data Availability

all data are available upon request

